# Dietary Intake, Handgrip Strength, and Elevated Blood Pressure Among Community-Dwelling Older Adults in Peri-Urban Ghana: A Cross-sectional Study

**DOI:** 10.64898/2026.10.13.26365389

**Authors:** Matilda Anim-Fofie, Gloria E. Otoo, Obed Akwaa Harrison, Jolene M. Azagba-Nyako, Matilda Steiner-Asiedu, Firibu K. Saalia

## Abstract

**Background:** Aging is associated with physiological, biological, socioeconomic and psychological changes that may increase vulnerability to poor dietary intake, reduced muscle strength and elevated blood pressure. In Ghana, evidence on dietary intake in relation to functional and cardiometabolic health among community-dwelling older adults remains limited. This cross-sectional study assessed dietary intake, handgrip strength (HGS) and elevated blood pressure (BP), and examined associations between diet quality indicators and these health outcomes among older adults in five peri-urban communities in the La Nkwantanang-Madina Municipality, Ghana.

**Methods:** Data were collected from 200 community-dwelling adults aged 60 years and above. Dietary intake was assessed using a multiple-pass 24-hour dietary recall, while physical activity, HGS and BP were measured using standardized procedures. Categorical variables were summarized as frequencies and percentages. Non-normally distributed dietary variables were compared between health outcome groups using the Mann-Whitney U test. Adjusted binary logistic regression was used to examine associations between diet quality indicators and HGS or elevated BP.

**Results:** The mean age of participants was 69.8 ± 7.3 years. More than half did not meet the dietary reference intake for protein, fat, fiber, magnesium, potassium and vitamin E, and no participant met the dietary reference intake for calcium. The prevalence of inadequate minimum dietary diversity (MDD) and inadequate mean adequacy ratio (MAR) exceeded 50%. Energy, protein, carbohydrate and selenium intake differed significantly by HGS status (p < 0.05). Mean adequacy ratio differed significantly by HGS status (p = 0.021) and BP status (p = 0.016). In adjusted models, each one-unit increase in MDD was associated with lower odds of elevated BP (AOR = 0.68; 95% CI: 0.46–1.00; p = 0.05), while each one-unit increase in MAR was associated with higher odds of normal HGS (AOR = 8.77; 95% CI: 1.33–64.08; p = 0.034).

**Conclusion:** Dietary inadequacy was common among older adults in this peri-urban Ghanaian setting. Better diet quality indicators were associated with normal HGS and lower odds of elevated BP. These findings support the need for nutrition-focused strategies and further longitudinal or intervention studies to clarify the role of dietary intake in functional and cardiometabolic health among older adults.

## Introduction

The United Nations (UN) classifies persons aged 60 years and above as older adults or belonging to the older population (1). Using 2021 as the baseline year, it is estimated that there would be a two-fold increase of the older population from 1.1 billion to 2.1 billion by the middle of the 21^st^ century and low- and middle-income countries (LMICs) would be home to two-thirds of this estimation (2). In sub–Saharan African (SSA) countries, older adults remain a smaller proportion of the population; however, their growth is projected to be four times that of developed countries by 2050 (3). Ghana has equally recorded a sharp rise in her older population by a near 10-fold increase from 213,477 in 1960 to about 2 million in 2021, as indicated in the report on the Population and Housing Census conducted in 2021 (4). Decline in infant mortality and an increase in life expectancy due to improved health care delivery are significant contributors to the global demographics of aging.

The physiological, biological, psychological and socioeconomic changes that accompany aging partly presents aging as a central risk factor for malnutrition and poor health outcomes through an interconnected pathway (5, 6). Furthermore, in LMICs, widespread poverty and food insecurity in the years preceding old age remain prominent among the causes of malnutrition and poor health outcomes in older adults. Unfortunately, the situation mostly remains unchanged during geriatric years.

The WHO describes undernutrition among older adults as common, with a prevalence ranging from 1.3% to 47.8% among community-dwelling older adults (7). The following studies conducted in Ghana have reported varying prevalence of nutritional status among older adults. A 10% underweight, 51.7% normal body mass index, 21.7% overweight, 16.6% obesity and 53% poor dietary pattern was found in four peri-urban communities (8). Blankson and Hall (9) reported 41% underweight, 42% normal body mass index, 15% overweight and 2% obesity among older adult women. Among a sample of urban dwellers, (10) reported 18.0% underweight, 60.5% normal body mass index and 21.5% overweight. Biochemical indicators of vitamins C (86.1%) and E (81.3%) were below recommended levels in the majority of participants in a study by Apprey, Kalog (11). Insufficient intake of both macro and micro nutrients was evident among older adults in the Effutu Municipality (12).

Some health problems common among older adults are hypertension and loss of muscle strength. Longevity increases the risk of developing hypertension by more than 90% (13). Among the top five causes of morbidity in Ghana is hypertension (14). Handgrip strength (HGS), a measure of muscle strength, declined with age in a nationally representative WHO survey conducted in Ghana (15). To the aging population, HGS has become relevant due to its connection to the concept of sarcopenia and the degree of independence to carry out certain day-to-day activities on their own, usually referred to as activities of daily living (ADL), such as self-care, mobility, bathing, toileting, feeding oneself, preparing and cooking meals (16, 17). Malnutrition increases vulnerability to muscle decline and chronic diseases, especially among older adults. The role of nutrition in attenuating the impact of aging on HGS and hypertension is well documented (18). Muscle synthesis requires the consumption of both high-quality and adequate protein and energy. Unfortunately, reduced food intake associated with aging makes it challenging to meet nutrient intake.

Although previous studies in Ghana have described nutritional status and selected health outcomes among older adults, fewer studies have jointly examined nutrient adequacy, dietary diversity, HGS and elevated BP among community-dwelling older adults in peri-urban settings. This evidence gap is important because peri-urban communities may experience rapid changes in food access, livelihoods and health risks. Therefore, this cross-sectional study assessed the relationship between dietary intake and selected health outcomes among older adults in peri-urban communities of the La Nkwantanang-Madina Municipality in the Greater Accra Region, Ghana. Specifically, the study compared dietary intake by HGS and BP status and examined whether diet quality indicators were associated with these outcomes.

## Materials and Methods

### Study design and setting

This cross-sectional survey served as phase I of an oral nutritional supplementation pilot study that included a baseline cross-sectional survey and a subsequent randomized controlled trial (RCT). The RCT is registered at www.clinicaltrials.gov with the registration number NCT06773975. The present manuscript reports only findings from the cross-sectional survey. The study was conducted in five peri-urban communities (Ayi Mensah, Kweiman, Danfa, Adoteiman and Otinibi) of the La Nkwantanang-Madina Municipality in the Greater Accra Region, Ghana, from November 2024 to January 2025. La Nkwantanang-Madina Municipality is predominantly urban, with approximately 16% of its population residing in peri-urban areas.

### Study population and eligibility criteria

Participants were recruited based on eligibility criteria for the RCT. Participants who consented to participate were to be 60 years and above, not allergic to the ingredients of the oral nutritional supplement, non-consumers of oral nutritional supplements, without sickle cell disease, terminal illness, clinical depression, impaired speech, hearing, vision or cognition.

### Sampling procedure and sample size

The five peri-urban communities were conveniently selected based on the following reasons. There were no studies published on the nutritional status of older adults in the La Nkwantanang-Madina Municipality. Furthermore, there is an apparent threat to food security and vulnerability to malnutrition due to rapid urbanization of the peri-urban communities and consequent loss of farmland. Moreover, proximity of the area to the University of Ghana was favorable in maximizing the limited resources allocated to the study.

The sample size was determined because of an earlier decision to conduct the RCT for 200 participants. Based on the following assumptions using the G*Power software: Cohen’s effect size of 0.3, a statistical power of 80%, a 5% level of significance, fixed effect, main effect interaction for repeated measures, a sample size of 90 participants was generated for each of the two arms of the RCT. An estimated attrition rate of 10% resulted in a total sample size of 200 participants. A census of individuals aged 60 years and above living in the communities was conducted to enumerate eligible participants. Proportional allocation per community was used to randomly select 200 participants from 276 eligible individuals enumerated at the census.

### Data collection and variables

#### Sociodemographic characteristics

A pretested semi-structured questionnaire was used to solicit information on sociodemographic characteristics.

#### Dietary intake

The multiple-pass 24-hour dietary recall method was used to collect information on all foods and drinks consumed the previous day. Food models and household food measures were shown to participants to improve estimation of portion size. Portion sizes of foods and drinks were converted into grams using a compiled Excel sheet designed by the Department of Nutrition and Food Science, University of Ghana that has the weight of foods. The weights of the foods were entered in the Ghana Foods Nutrient Database Version 1.0 software for analysis.

The preferred dietary reference intake (DRI) for populations is the estimated average requirement (EAR), which represents the average daily nutrient intake estimated to meet the requirements of half of healthy individuals in a group. Adequate intake was used for nutrients without an EAR, and the acceptable macronutrient distribution range (AMDR) was used for fat. Sex-specific EAR values for protein were estimated as 0.66 g/kg body weight (19). Reference body weights of 70 kg for men and 57.5 kg for women were used for protein. To assess dietary diversity, foods from the multiple-pass 24-hour dietary recall were grouped into the 10 food groups used in the Minimum Dietary Diversity for Women (MDD-W) indicator by the Food and Agriculture Organization (20). In this study, MDD-W food groups were used as a practical proxy for dietary diversity among older adults; however, the indicator was not originally developed specifically for older adults or mixed-sex geriatric populations. Mean adequacy ratio was calculated as the average of nutrient adequacy ratios (NARs) for nutrients identified from the literature as relevant to HGS and BP. Separate MAR scores were calculated for HGS and BP. Based on the literature, protein, carbohydrate, iron, calcium, leucine, selenium and vitamins C and E were selected for HGS, while carbohydrate, fat, fiber, cholesterol, calcium, magnesium, potassium, sodium, selenium and vitamins C and E were selected for BP (21–24).

#### Physical activity

An adapted physical activity scale for the elderly (PASE) by Washburn, Smith (25) to include activities common with older adults in Ghana was used. The PASE collects information on self-reported occupational, household and leisure activities that took place 7 days before the interview and their frequency of occurrence. PASE scores were calculated from activity weight and activity frequency.

#### Handgrip strength

An electronic dynamometer (model number: EH108) held at the elbow and bent at a 90-degree angle was used to measure the HGS of the dominant arm. The dynamometer was held for 3 seconds and HGS was measured to the nearest 0.1 kg and repeated within a minute interval. The mean of 3 readings was recorded as handgrip strength.

#### Blood pressure

Participants were seated in a rested position in a chair, with their backs against the chair and their legs uncrossed. No tight clothing was worn on the upper left arm, which rested on a table. After a 5-minute rest, a digital OMRON automatic sphygmomanometer Model: M2 (HEM-7121-E) was used to take two measurements from the upper left arm within a 2-minute interval from each reading. A third reading was recorded if the difference between the first and second readings was greater than 10 mmHg. The mean of two readings to the nearest mmHg was recorded as the blood pressure.

### Operational definitions of variables/indicators

#### Dietary intake

Nutrient intakes were classified as below the DRI and met the DRI when intake was ≥ the DRI. For sodium, it is more beneficial not to exceed the DRI. Participants with intake ≤ DRI were classified as met the DRI and intake > DRI as below the DRI. Participants whose intake were within the AMDR were categorized as met the DRI for fat and outside of the range as below the DRI. Body weight measurement is included in the calculation of the estimated energy requirement (EER). However, body weight measurements were unavailable for the cross-sectional survey, making the evaluation of EER impossible. No official EAR values exist for cholesterol and leucine. Therefore, categorization of participants into below or met the DRI for energy/calories, cholestereol and leucine were omitted.

Two indicators, the minimum dietary diversity score (MDDS) and the mean adequacy ratio (MAR) were used to define diet quality. Participants who consumed <5 food groups had inadequate MDDS and a consumption of ≥5 groups is adequate MDDS. Participants were categorized into a No response if they did not consume food from a food group and a Yes response if they did. An MAR score of ≥ 0.75 is considered adequate and < 0.75 is inadequate (26).

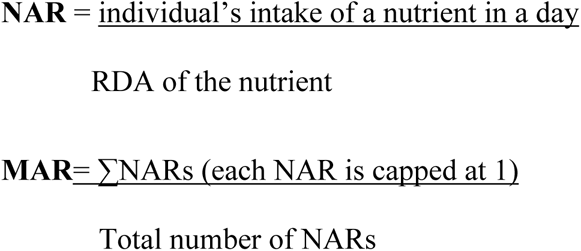

#### Health outcomes

The health outcomes were categorized as weak/normal HGS and normal/elevated BP. The cut-offs recommended by the European Working Group on Sarcopenia in Older People 2 (EWGSOP2) were used to classify weak HGS as <27 kg for males and <16 kg for females, and normal HGS as ≥27 kg for males and ≥16 kg for females. Because BP was measured on one day only, the outcome is described as elevated BP rather than diagnosed hypertension. Systolic BP ≥140 mmHg and/or diastolic BP ≥90 mmHg was classified as elevated BP while systolic BP <140 mmHg and or diastolic BP <90 mmHg was classified as normal BP, consistent with WHO recommendations (27).

### Quality control and data management

The questionnaires were pretested among 20 participants from two peri-urban communities in the Ga West Municipality of the Greater Accra Region. The data collectors were trained in interviewing/communication skills and use of the instruments for data collection. After each day’s data collection, all questionnaires were checked for completeness before being submitted. Only the principal investigator had access to copies of signed informed consent forms. Data for analyses did not bear the names of participants. All participants were given unique ID numbers to conceal their identity. All dietary recalls that had caloric intake below 1000 calories or above 4000 calories were reviewed to verify the reported intake.

### Statistical analysis

IBM SPSS version 27 was used for analyses. Statistical significance was set at p < 0.05. Categorical variables were presented as frequencies and percentages. Continuous variables were assessed for normality. Normally distributed variables were presented as means ± standard deviations, while non-normally distributed variables were presented as medians and inter-quartile ranges (IQRs). The Mann-Whitney U test was used to compare dietary intake between categories of health outcomes. Associations between sociodemographic characteristics and health outcomes were examined using bivariate analyses of the Chi square test of independence or the alternative Fisher’s exact test for expected cell count below five. Adjusted binary logistic regression models were used to examine factors associated with HGS and BP status. Minimum dietary diversity and MAR were included as dietary predictors because foods are consumed as combinations of nutrients rather than as isolated nutrients. Age, PASE score and use of anti-hypertensive drug were included as covariates for BP status, while age, sex and PASE score were included as covariates for HGS status. Sociodemographic characteristics that were significantly associated with health outcomes in bivariate analyses were also included as covariates. Minimum dietary diversity score and MAR were treated as continuous independent variables in their respective regression models.

### Ethical considerations

This study was conducted in accordance with the principles of the Helsinki Declaration on research involving human participants. Ethical approval was obtained from the Ethics Committee of the College of Basic and Applied Science (ECBAS) of the University of Ghana (UG-ECBAS 113/23-24). Before the commencement of data collection, an explanation of the study protocol was given to all participants. Participants gave their consent by signing or thumbprinting the written informed consent or a witness signed in their stead if the participant could not read the form.

## Results

### Sociodemographic characteristics of participants

**Table 1** describes the sociodemographic characteristics of participants. The mean age of participants was 69.8 ± 7.3 years. More than half were female, Christian, had primary/middle school education, had dependents and had a caretaker. Nearly all participants (97.0%) had a source of livelihood. The largest ethnic group was Ga/Dangme (33.5%), and slightly more than half of participants were employed (50.5%). In bivariate analyses (**Table 2**), BP status differed significantly by religious affiliation (p = 0.013) and dependent status (p = 0.035). Elevated BP was more common among participants classified as belonging to other religious groups than among Christians, and among participants without dependents than among those with dependents. HGS status differed significantly by employment status (p = 0.024) and source of livelihood (p = 0.009), with weak HGS more common among unemployed participants and among those without a source of livelihood. These significant bivariate associations were included in the binary logistic regression as covariates to confirm their true significant association with the health outcomes.

**Table 1:** Sociodemographic characteristics of participants.

| **Sociodemographic characteristics** | **N (%)** |
| --- | --- |
| **Sex** |  |
| Female | 142 (71.0) |
| **Marital status** |  |
| Single | 109 (54.5) |
| Married/cohabit | 91(45.5) |
| **Level of Education** |  |
| None | 53 (26.5) |
| Primary/Middle | 105 (52.5) |
| Secondary/Higher | 42 (21.0) |
| **Religion** |  |
| Christian | 168 (84.0) |
| Others | 32 (16.0) |
| **Ethnicity** |  |
| Ga/Dangme | 67 (33.5) |
| Ewe | 49 (24.5) |
| Akan | 49 (24.5) |
| Northern | 35 (17.5) |

**Table 1 continued: Sociodemographic characteristics of participants**
| **Sociodemographic characteristics** | **N (%)** |
| --- | --- |
| **Has dependents** |  |
| Yes | 145 (72.5) |
| **Has a caretaker** |  |
| Yes | 127 (63.5) |
| **Lives alone** |  |
| Yes | 24 (12.0) |
| **Employed** |  |
| Yes | 101 (50.5) |
| **Has a source of livelihood** |  |
| Yes | 194 (97.0) |

**Table 2:** Bivariate analysis between sociodemographic characteristics and health outcomes.

| Sociodemographic characteristics | HGS |  |  | BP |  |  |
| --- | --- | --- | --- | --- | --- | --- |
|  | Weak | Normal | p- | Normal | Elevated | p- |
|  | HGS | HGS | value | BP | BP | value |
|  | N (%) | N (%) |  | N (%) | N (%) |  |
| Sex |  |  |  |  |  |  |
| Male | 31 (53.4) | 27 (46.6) | 0.208 | 33 (56.9) | 25 (43.1) | 0.082 |
| Female | 62 (43.7) | 80 (56.3) |  | 99 (69.7) | 43 (30.3) |  |
| Marital status |  |  |  |  |  |  |
| Single | 48 (44.0) | 61 (56.0) | 0.584 | 70 (64.2) | 39 (35.8) | 0.561 |
| Married/cohabit | 45 (49.5) | 46 (50.5) |  | 62 (68.1) | 29 (31.9) |  |
| Level of Education |  |  |  |  |  |  |
| None | 31 (58.5) | 22 (41.5) | 0.111 | 31 (58.5) | 22 (41.5) | 0.287 |
| Primary/Middle | 43 (41.0) | 62 (59.0) |  | 70 (66.7) | 35 (33.3) |  |
| Secondary/Higher | 19 (45.2) | 23 (54.8) |  | 31 (73.8) | 11 (26.2) |  |
| Religion |  |  |  |  |  |  |
| Christian | 76 (45.2) | 92 (54.8) | 0.412 | 117(88.6) | 17 (11.4) | 0.013 |
| Others | 17 (53.1) | 15 (46.9) |  | 15 (46.9) | 51 (53.1) |  |
| Ethnicity |  |  |  |  |  |  |
| Ga/Dangme | 29 (43.3) | 38 (56.7) | 0.544 | 44 (65.7) | 23 (34.3) |  |
| Ewe | 21 (42.9) | 28 (57.1) |  | 35 (71.4) | 14 (28.6) | 0.203 |
| Akan | 23 (46.9) | 26 (53.1) |  | 35 (71.4) | 14 (28.6) |  |
| Northern | 20 (57.1) | 15 (42.9) |  | 18 (51.4) | 17 (48.6) |  |
|  | Weak | Normal | p- | Normal | Elevated | p- |
|  | HGS | HGS | value | BP | BP | value |
|  | N (%) | N (%) |  | N (%) | N (%) |  |
| **Has dependents** |  |  |  |  |  |  |
| No | 29 (52.7) | 26 (47.3) | 0.277 | 30 (54.5) | 25 (45.5) | **0.035** |
| Yes | 64 (44.1) | 81 (55.9) |  | 102(70.3) | 43 (29.7) |  |
| **Has a caretaker** |  |  |  |  |  |  |
| No | 33 (45.2) | 40 (54.8) | 0.781 | 43 (58.9) | 30 (41.1) | 0.108 |
| Yes | 60 (47.2) | 67 (52.8) |  | 89 (70.1) | 38 (29.9) |  |
| **Lives alone** |  |  |  |  |  |  |
| No | 82 (46.6) | 94 (53.4) | 0.944 | 116(65.9) | 60 (34.1) | 0.941 |
| Yes | 11 (45.8) | 13 (54.2) |  | 16 (66.7) | 8 (33.3) |  |
| **Employed** |  |  |  |  |  |  |
| No | 54 (54.5) | 45 (45.5) | **0.024** | 64 (64.6) | 35 (35.4) | 0.689 |
| Yes | 39 (38.6) | 62 (61.4) |  | 68 (67.3) | 33 (32.7) |  |
| **Has a source of livelihood** |  |  |  |  |  |  |
| No |  |  |  |  |  |  |
| Yes | 6 (100.0) | 0 (0.0) | **0.009** | 4 (66.7) | 2 (33.3) | 1.000 |
|  | 87 (44.8) | 107(55.2) |  | 128(66.0) | 66 (34.0) |  |

### Health outcomes of participants

The prevalence of weak HGS was 46.5%, while elevated BP was observed in 34.0% of participants.

### Dietary assessment of participants

**Table 3** summarizes the nutrient intake of participants. More than half of the participants had intake below the DRI for protein, fat, fiber, magnesium, potassium and vitamin E. None met the DRI for calcium. In **Figure 1**, except for 3 food groups (grains, white roots, tubers and plantains; meat, poultry and fish; other vegetables), majority of the participants (ranging from 80-91%) did not consume any of the other 7 food groups. More than half of participants were classified as being inadequate for each of the indicators for diet quality (Figure 2).

**Figure 1:**
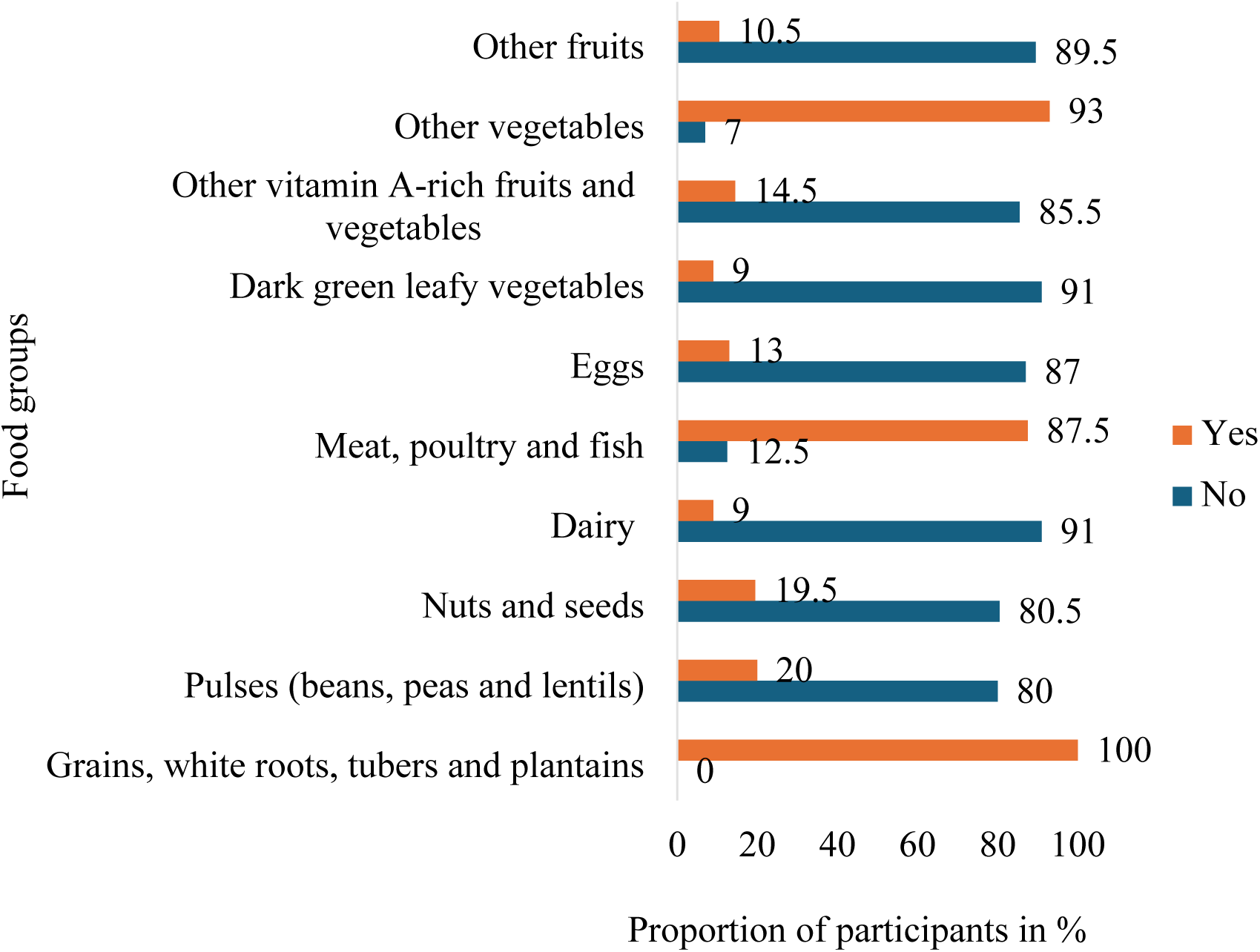
Consumption of food groups.

**Figure 2:**
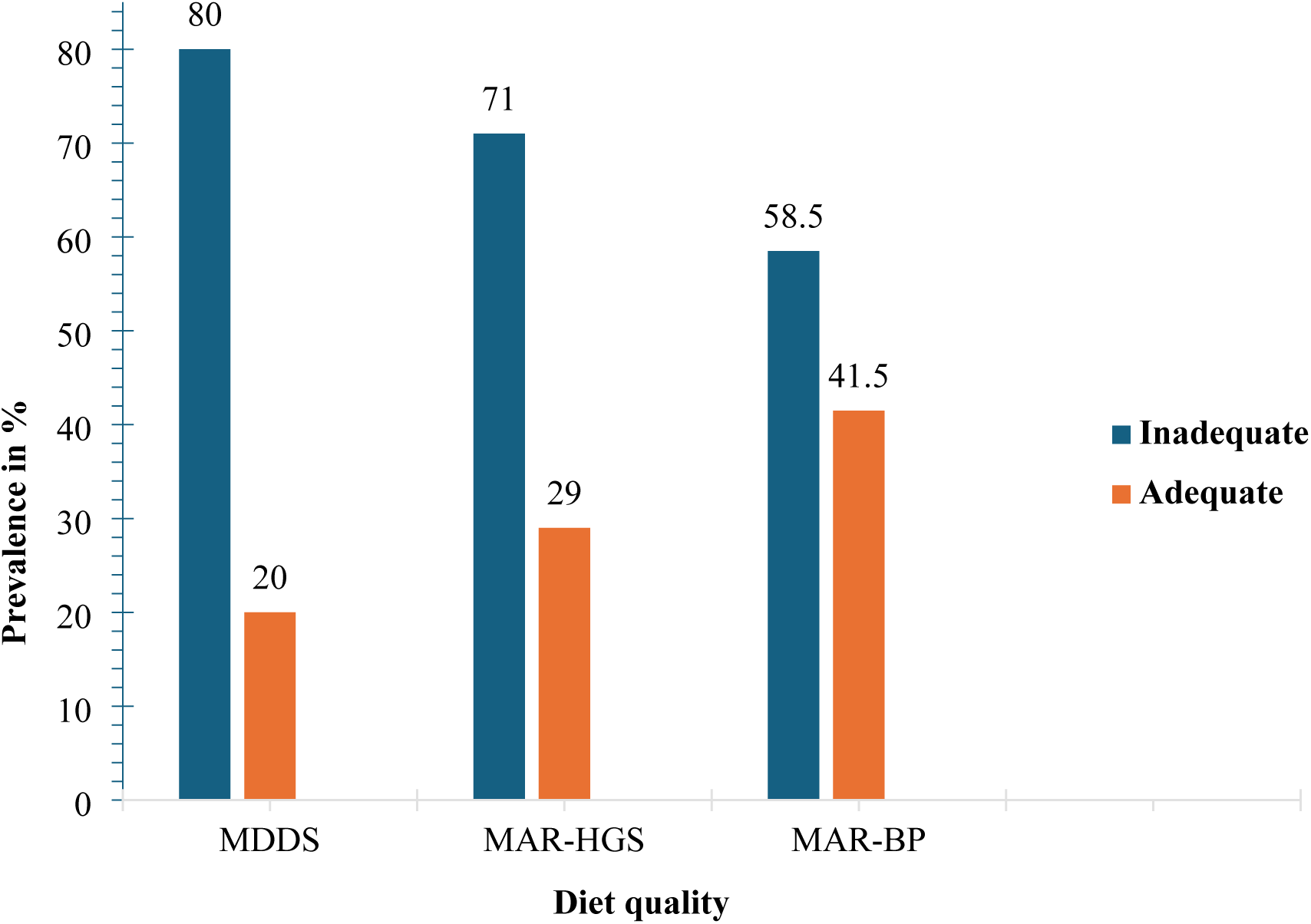
Prevalence of diet quality.

**Table 3:**
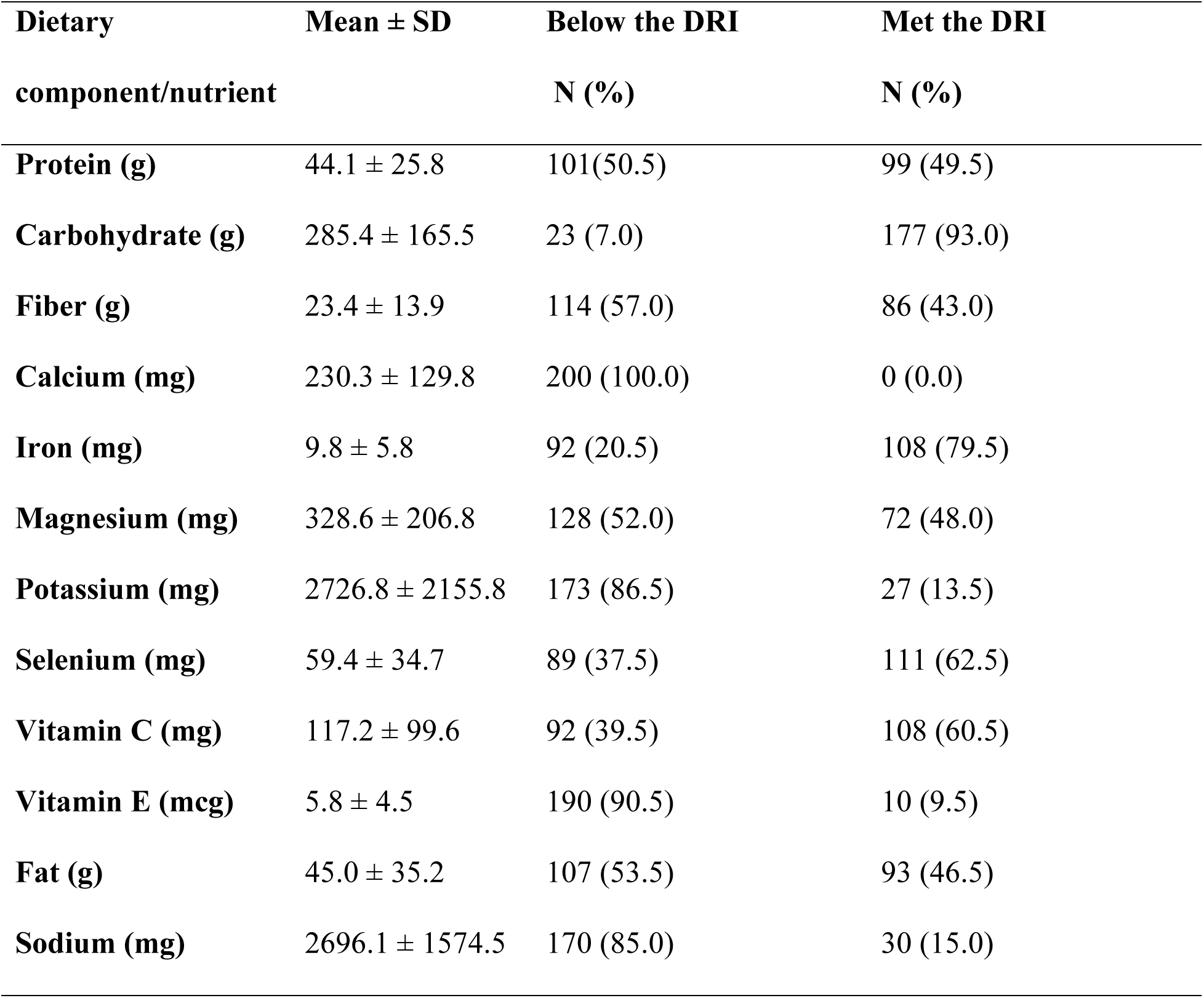
Prevalence of nutrient intake of participants.

| **Dietary**<br>**component/nutrient** | **Mean ± SD** | **Below the DRI**<br>**N (%)** | **Met the DRI**<br>**N (%)** |
| --- | --- | --- | --- |
| **Protein (g)** | 44.1 ± 25.8 | 101(50.5) | 99 (49.5) |
| **Carbohydrate (g)** | 285.4 ± 165.5 | 23 (7.0) | 177 (93.0) |
| **Fiber (g)** | 23.4 ± 13.9 | 114 (57.0) | 86 (43.0) |
| **Calcium (mg)** | 230.3 ± 129.8 | 200 (100.0) | 0 (0.0) |
| **Iron (mg)** | 9.8 ± 5.8 | 92 (20.5) | 108 (79.5) |
| **Magnesium (mg)** | 328.6 ± 206.8 | 128 (52.0) | 72 (48.0) |
| **Potassium (mg)** | 2726.8 ± 2155.8 | 173 (86.5) | 27 (13.5) |
| **Selenium (mg)** | 59.4 ± 34.7 | 89 (37.5) | 111 (62.5) |
| **Vitamin C (mg)** | 117.2 ± 99.6 | 92 (39.5) | 108 (60.5) |
| **Vitamin E (mcg)** | 5.8 ± 4.5 | 190 (90.5) | 10 (9.5) |
| **Fat (g)** | 45.0 ± 35.2 | 107 (53.5) | 93 (46.5) |
| **Sodium (mg)** | 2696.1 ± 1574.5 | 170 (85.0) | 30 (15.0) |

### Significant relationships between dietary intake and health outcomes

As shown in **Table 4**, participants with normal HGS had significantly higher intake of energy, protein, carbohydrate and selenium than those with weak HGS (p < 0.05). Mean adequacy ratio was also significantly higher among participants with normal HGS (p = 0.021). For BP status, MAR was significantly higher among participants with normal BP than among those with elevated BP (**Table 5**; p = 0.016). In the adjusted regression model for HGS (**Table 6**), MAR was associated with higher odds of normal HGS (AOR = 8.65; 95% CI: 1.16-64.42; p = 0.035). In the adjusted regression model for BP (**Table 7**), participants in the “other religion” category had higher odds of elevated BP than Christians (AOR = 3.49; 95% CI: 1.49–8.15; p = 0.004). These associations should be interpreted as exploratory because of the cross-sectional design and sparse data in some categories.

**Table 4:**
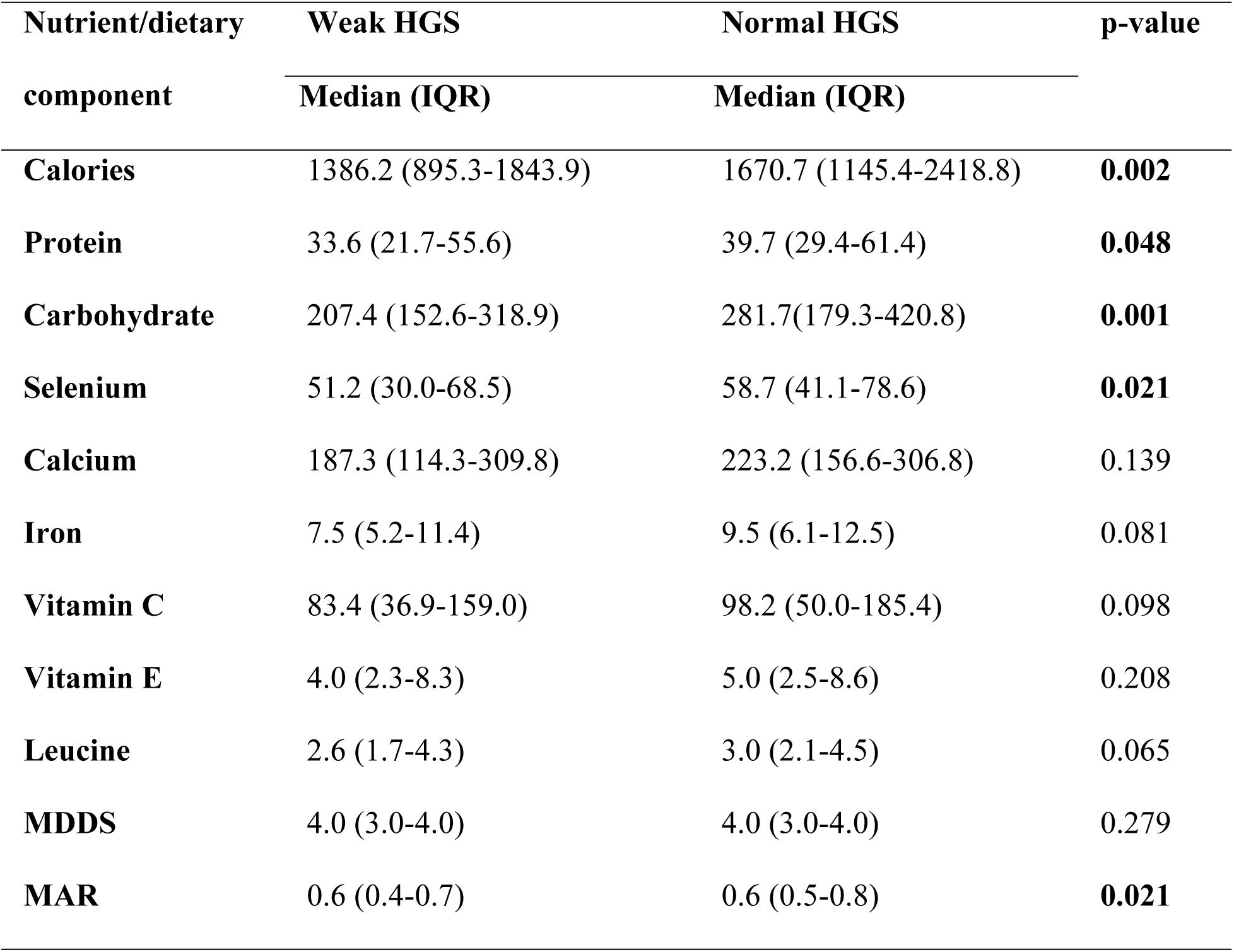
Comparison of dietary intake between weak and normal HGS.

| Nutrient/dietary<br>component | Weak HGS<br>Median (IQR) | Normal HGS<br>Median (IQR) | p-value |
| --- | --- | --- | --- |
| **Calories** | 1386.2 (895.3-1843.9) | 1670.7 (1145.4-2418.8) | **0.002** |
| **Protein** | 33.6 (21.7-55.6) | 39.7 (29.4-61.4) | **0.048** |
| **Carbohydrate** | 207.4 (152.6-318.9) | 281.7(179.3-420.8) | **0.001** |
| **Selenium** | 51.2 (30.0-68.5) | 58.7 (41.1-78.6) | **0.021** |
| **Calcium** | 187.3 (114.3-309.8) | 223.2 (156.6-306.8) | 0.139 |
| **Iron** | 7.5 (5.2-11.4) | 9.5 (6.1-12.5) | 0.081 |
| **Vitamin C** | 83.4 (36.9-159.0) | 98.2 (50.0-185.4) | 0.098 |
| **Vitamin E** | 4.0 (2.3-8.3) | 5.0 (2.5-8.6) | 0.208 |
| **Leucine** | 2.6 (1.7-4.3) | 3.0 (2.1-4.5) | 0.065 |
| **MDDS** | 4.0 (3.0-4.0) | 4.0 (3.0-4.0) | 0.279 |
| **MAR** | 0.6 (0.4-0.7) | 0.6 (0.5-0.8) | **0.021** |

**Table 5:** Comparison of dietary intake between normal and elevated BP.

| **Nutrient/dietary component** | **Normal BP** | **Elevated BP** | **p-value** |
| --- | --- | --- | --- |
|  | **Median (IQR)** | **Median (IQR)** |  |
| **Calories** | 1469.6 (1028.4- 2175.2) | 1600.7 (1032.8-2470.6) | 0.512 |
| **Fat** | 35.9 (18.9- 62.9) | 34.1 (20.8-62.3) | 0.981 |
| **Carbohydrate** | 238.7 (165.8- 350.6) | 250.9 (163.2-418.1) | 0.432 |
| **Fiber** | 20.0 (12.6- 32.8) | 19.7 (12.9-34.9) | 0.922 |
| **Cholesterol** | 39.5 (22.8- 94.7) | 54.2 (28.1-6106.0) | 0.350 |
| **Calcium** | 203.5 (137.8- 299.8) | 194.5 (148.2-328.5) | 0.798 |
| **Magnesium** | 278.6 (169.1- 436.6) | 284.9 (192.6-471.7) | 0.831 |
| **Potassium** | 2172.5 (1242.1- 306.0) | 2034.6 (1084.8-4451.5) | 0.904 |
| **Sodium** | 2321.8 (1555.4- 3172.1) | 2661.5(1678.5-3878.5) | 0.185 |
| **Selenium** | 53.4 (36.9- 72.4) | 52.4(34.36-79.1) | 0.494 |
| **Vitamin C** | 94.6 (42.9-171.2) | 81.9 (39.0-166.1) | 0.661 |
| **Vitamin E** | 4.8 (2.4- 8.6) | 4.2 (2.0-7.6) | 0.326 |
| **MDDS** | 4.0 (3.0-4.0) | 3.0 (3.0-4.0) | 0.164 |
| **MAR** | 0.7 (0.6-0.8) | 0.6 (0.5-0.8) | **0.016** |

**Table 6:** Predictors of HGS among participants.

| **Predictors** | **AOR** | **95% CI** | **p-value** |
| --- | --- | --- | --- |
| **Age of participant** | 0.95 | 0.91-1.00 | 0.213 |
| **PASE** | 0.99 | 0.99-1.00 | 0.384 |
| **Sex of participant** |  |  |  |
| Male (Ref) | 1.00 |  |  |
| Female | 0.58 | 0.29-1.16 | 0.120 |
| **Employed** |  |  |  |
| Yes (Ref) | 1.00 | 0.31-1.15 |  |
| No | 0.60 |  | 0.120 |
| **Has a source of livelihood** |  |  |  |
| Yes (Ref) | 1.00 | 0.00-. |  |
| No | 0.00 |  | 0.999 |
| **MAR** | 8.65 | 1.16-64.42 | **0.035** |
| **MDDS** | 1.03 | 0.73-1.46 | 0.84 |

**Table 7:** Predictors of BP among participants.

| **Predictors** | **AOR** | **95% CI** | **p-value** |
| --- | --- | --- | --- |
| **Age of participant** | 0.95 | 0.91-1.00 | 0.064 |
| **PASE** | 1.00 | 0.99-1.00 | 0.384 |
| **Religion** |  |  |  |
| Christian (Ref) | 1.00 |  |  |
| Other | 3.49 | 1.49-8.15 | **0.004** |
| **Have dependents** |  |  |  |
| Yes (Ref) | 1.00 |  |  |
| No | 2.19 | 1.01-4.13 | **0.026** |
| **Use of hypertensive drug** |  |  |  |
| Yes (Ref) | 1.00 |  |  |
| No | 1.13 | 0.58-2.20 | 0.710 |
| **MAR** | 1.78 | 0.24-12.89 | 0.609 |
| **MDDS** | 0.68 | 0.46-1.00 | 0.064 |

## Discussion

This cross-sectional study assessed dietary intake, diet quality indicators, HGS and elevated BP among community-dwelling older adults in peri-urban Ghana. The findings showed a high prevalence of dietary inadequacy, substantial prevalence of weak HGS and elevated BP, and associations between selected diet quality indicators and health outcomes. The study contributes to the limited evidence on nutrition, functional health and cardiometabolic risk among older adults in peri-urban Ghana. However, because the design was cross-sectional, the findings should be interpreted as associations rather than evidence of causal relationships.

The mean age of the participants in this study was same as the mean age of older adult participants of a cross-sectional survey in the Sunyani municipality in Ghana (11). Apprey, Kalog (11), (12) found more females than males in their study among older adults in Ghana. More than half of the participants had formal education, similar to findings from Amfo-Antiri, Agyapong (28) among older adult respondents in the Kwahu South District of the Eastern Region, Ghana. A little more than half (50.5%) of the participants in this study were employed, higher than the national average for older adults (37.5%) according to the 2021 population and housing census (PHC) (4). The proportion (12.0%) of participants who lived alone was lower than older adults in the general population (17%) of the 2021 PHC.

Both macro and micronutrient deficiencies are common among the older population and were evident from this cross-sectional survey. Similar to the findings by Hayfron, Annan (12) from older adults in the Effutu Municipality of the Central region, Ghana, more than half of participants did not meet their dietary intake for protein, fat, fiber, magnesium, potassium, vitamin E. None met the requirement for calcium. Oldewage-Theron, Samuel (29) reported inadequate intake for energy, protein, iron, zinc, vitamins A, C and E in 25% of participants in South Africa. Except for the grains, white roots, tubers and plantains food group where all participants consumed the previous day, consumption of each of the other 9 food groups ranged from 7-93%. The prevalence of inadequate MDD (80%) was higher than (53.3%) among participants from peri urban communities of the Ga West municipality in the Greater Accra of Ghana (8) and lower than that of a study across six provinces in Sri Lanka where none of the three indicators used to measure dietary diversity met the optimum level defined as 7 out of 12 food groups (30).

This study corroborates existing evidence that dietary diversity remains a challenge in LMIC due to the limitation in the consumption of micronutrients because diets are typically starchy staples that contain little or no animal products and a variety of fruits and vegetables (31).

Handgrip strength (HGS) has gained recognition as a supportive indicator in the assessment of nutritional status (32). Weak HGS is considered a primary parameter for sarcopenia and in the assessment of muscle function, muscle strength is said to be the most reliable measure (33). A lack of uniformity in the methods for assessing the two health outcomes (HGS and BP) makes comparison of findings challenging (34). There is heterogeneity such as the age of the study population and cut-offs. Moreover, there are limited guidelines and studies on HGS from LMIC compared to high-income countries (35, 36).

The prevalence of weak HGS was a little less than half (46.5%) of the study population. A study among rural community-dwelling individuals aged 50 years and above in the Garu-Tempane District in the Upper East Region of Ghana (36) and the nationally representative WHO-SAGE Wave 1 Survey in Ghana (15) did not classify participants as having weak or normal HGS; however, both reported higher mean HGS than those found in this study (25.6 kg ± 10.3 kg and 17.3 kg ± 7.1 kg for males and females, respectively). The higher HGS in the two studies could be due to a younger study population than this study in LaNNMA as decline in HGS is strongly correlated with increasing chronological age (37).

Unlike this study, which reported overall weak HGS prevalence (46.5%), Wearing, Konings (38) presented a sex-specific prevalence of 44% among men and 53% among women aged ≥ 75 years. The prevalence in this study was higher than the 2017 and 2018 (32.3%) Korea National Health and Nutrition Examination Survey (KNHANES) among 3,075 individuals aged above 60 years (39). Using the same HGS cut-off as this study, the prevalence of weak HGS (27.3%) from a cross-sectional population-based Israeli Health and Nutrition Survey of older adults was lower than that recorded in this study (40).

The relatively low prevalence of elevated BP (34%) in this study could be attributed to the younger mean age of the study population. Elevated BP is associated with increasing age, with a prevalence of 27% among persons aged below 60 years to 74% among those above 80 years (41). Apprey, Kalog (11) found a prevalence 19% among 375 participants with a mean age same as that of this study in a cross-sectional survey in the Sunyani municipality in Ghana. A higher prevalence of 50% was found among 3,575 participants aged 50 years and above in Ghana (42), possibly due to the larger sample size and diverse age distribution as a nationally representative survey.

Several factors influence the health outcomes of older adults. These include dietary intake, food insecurity, physical activity, socioeconomic conditions, chronic diseases, the natural progression of aging and these may generally differ across study populations.

Studies on the relationship between dietary intake and health outcomes in older adults have produced mixed findings (43). Protein mostly dominates discussions relating to the development of sarcopenia; however, micronutrients and other components of the diet have a significant contribution in the prevention and treatment of sarcopenia, and their role in muscle function and metabolism (44). Older adults who consume diverse diets suffer less decline in grip strength (45, 46).

Numerous studies have indicated a relationship between adequate consumption of both macro and micronutrients in addressing sarcopenia in older adults (47–49). Other components of the diet that have been found to modify the risk of sarcopenia are the total energy intake and the percentage contribution of energy from macronutrients.

Generally, in this study, there was a higher dietary intake among participants with normal HGS than those with weak HGS, although significant for only calories, protein, carbohydrate and selenium. In a study among community-dwelling older adults in Israel, Doyev et al. (2021) reported a significantly higher intake of calories, protein, carbohydrate and fat among participants with normal HGS than weak HGS. A cross-sectional study based on 2016 to 2019 KNHANES data showed a significantly higher protein, carbohydrate and fat intake for normal HGS among women, but only significant for caloric intake among men (50). A large general-population cohort study, the UK Biobank found a significantly negative association between carbohydrate intake and HGS. Beaudart, Locquet (23) and Park, Ham (24) reported a negative association between sarcopenia and total energy, protein and carbohydrates in older adults. A weak but significantly positive correlation between calories, protein, polyunsaturated fatty acid (PUFA), fiber, vitamin C and HGS in the 7th KNHANES (KNHANES VII-1 in 2016) (51).

Dietary assessment alone may not be the only predictor of health outcomes in older adults. Other predictors include socioeconomic status, education, employment, level of physical activity, age, sex, presence of inflammation and general wellbeing (52–58).

After adjustment for physical activity, age and sex, MAR was the only dietary indicator significantly associated with normal HGS. Each one-unit increase in MAR was associated with higher odds of normal HGS, suggesting that overall nutrient adequacy may be relevant to functional status in this population. However, the confidence interval was wide, indicating imprecision in the estimate. This finding is broadly consistent with the Singapore Multi-Ethnic Cohort study, which reported higher HGS among older participants with better diet quality. Huang, Shanmugam (59) found that the adequacy component of the dietary quality index–international was significantly associated with HGS.

Although nutrients such as sodium, potassium, calcium, magnesium, fiber and antioxidants have been linked to BP regulation, this study did not find significant differences in individual nutrient intake between BP groups. In contrast, an indicator for diet quality, the MAR, showed association with BP status: MAR was higher among participants with normal BP. This may reflect the importance of overall dietary patterns rather than single nutrients and corroborates an assertion by Huang, Shanmugam (59) that dietary quality may offer better insights into relationship between dietary intake and health outcomes than individual nutrients because dietary quality reflects overall dietary pattern. Nevertheless, the findings should be interpreted cautiously because the study design does not establish temporality, BP was measured on one day only, and residual confounding may remain. Dietary patterns such as Mediterranean, Nordic and Dietary Approaches to Stop Hypertension (DASH) diets are characterized by fruits, vegetables, whole grains, fish, nuts and dairy and have been associated with favorable cardiometabolic profiles (59–64). Fruits and vegetables also provide antioxidants and bioactive compounds that may influence inflammation and oxidative stress pathways (60, 61).

Physical activity is another modifiable factor relevant to muscle strength and elevated BP (62–64). The median PASE score of the study population was 102.3 (IQR: 63.8–147.5), which suggest low physical activity relative to total PASE score (approximately 793). PASE score did not differ significantly by BP status (p = 0.380), but it was significantly higher among participants with normal HGS than among those with weak HGS (p = 0.041). These findings suggest that physical activity may be relevant to functional health in this population, although causality cannot be inferred from these data.

The findings highlight a high burden of dietary inadequacy among older adults in this peri-urban Ghanaian setting. They also suggest that diet quality indicators may be relevant to functional and cardiometabolic health. Community-based nutrition strategies that promote dietary diversity and nutrient adequacy may therefore be useful components of healthy ageing programs. Such strategies could be combined with age-appropriate physical activity promotion. However, intervention studies are needed before concluding that improving diet quality will directly improve HGS or BP outcomes.

This study has limitations. First, the cross-sectional design limits causal inference and does not establish temporality. Second, the convenient selection of study communities limits the generalizability of findings beyond similar peri-urban settings. Third, HGS cut-offs were based on EWGSOP2 recommendations and are not specific to African populations. Fourth, dietary intake was assessed using a single 24-hour recall, which reflects recent intake and may not represent habitual individual intake. Fifth, MDD-W food groups were used as a proxy for dietary diversity, although the indicator was not originally designed for older adults or mixed-sex geriatric populations. Sixth, BP was measured on one day only; therefore, the outcome should be interpreted as elevated BP rather than diagnosed hypertension. Finally, residual confounding from factors such as medication use, comorbidities, food insecurity and socioeconomic conditions may remain. A major strength of the study is the assessment of HGS, an underreported functional outcome in studies among older adults in Ghana. The inclusion of dietary, functional and cardiometabolic measures also provides useful preliminary evidence for geriatric nutrition research in Ghana.

## Conclusion

Dietary assessment indicated a high prevalence of dietary inadequacy among older adults in the La Nkwantanang-Madina Municipality. Participants with normal HGS had higher intake of energy, protein, carbohydrate and selenium than those with weak HGS, and MAR was higher among participants with normal HGS and normal BP. Higher MAR was associated with higher odds of normal HGS. These findings suggest that dietary diversity and nutrient adequacy may be relevant to functional and cardiometabolic health among older adults in peri-urban Ghana. Further longitudinal and intervention studies are needed to confirm these associations and guide nutrition-focused healthy ageing interventions.

## Data Availability

All relevant data are within the manuscript and its Supporting Information files

## Acknowledgements

The authors express their gratitude to the elders and participants of the study communities for their cooperation. Special appreciation to the data collection team for their hard work and dedication.

## Authors’ contributions

Matilda Anim-Fofie developed the study protocol/proposal, supervised data collection, conducted the analysis, interpreted the data, and drafted the original manuscript. Gloria E. Otoo, Matilda Steiner-Asiedu, Jolene M. Azagba-Nyako, and Firibu K. Saalia refined the study protocol/proposal, provided academic supervision, reviewed and edited the manuscript. Obed Akwaa Harrison contributed to data interpretation, manuscript review and editing. Matilda Anim-Fofie, Gloria E. Otoo, Obed Akwaa Harrison, Jolene M. Azagba-Nyako, Matilda Steiner-Asiedu, and Firibu K. Saalia conceived the study and contributed to securing funding.

## Funding

This work was carried out with the aid of a grant from the O.R. Tambo Africa Research Chair Initiative as supported by University of Ghana; Ministry of Environment, Science and Technology (MEST); the National Research Foundation of South Africa (NRF); the Department of Science and Innovation of South Africa (DSI); the International Development Research Centre of Canada (IDRC); and the Oliver & Adelaide Tambo Foundation (OATF).

## Notes

### Competing Interest Statement

The authors have declared no competing interest.

### Clinical Trial

The trial is registered at www.clinicaltrials.gov with the registration number NCT06773975

### Author Declarations

Ethical approval was obtained from the Ethics Committee of the College of Basic and Applied Science (ECBAS) of the University of Ghana (UG-ECBAS 113/23-24).

## References

1. Kwong J. Social policies catalogue on population ageing: a rapid scoping review. Bangkok, Thailand: UNFPA Asia-Pacific Regional Office; 2020.

2. Tan MP. Healthcare for older people in lower and middle income countries. Age Ageing. 2022;51(4).

3. Velkoff VA, Kowal PR. Aging in sub-Saharan Africa: The changing demography of the region. Aging in sub-Saharan Africa: Recommendations for furthering research. 2006:55–91.

4. 2021 Population and Housing Census [press release]. 1st Ocotber 2022.

5. Besora-Moreno M, Llauradó E, Tarro L, Solà R. Social and Economic Factors and Malnutrition or the Risk of Malnutrition in the Elderly: A Systematic Review and Meta-Analysis of Observational Studies. Nutrients. 2020;12(3).

6. Shuremu M, Belachew T, Hassen K. Nutritional status and its associated factors among elderly people in Ilu Aba Bor Zone, Southwest Ethiopia: a community-based cross-sectional study. BMJ Open. 2023;13(1):e067787.

7. Organization WH. Integrated care for older people: guidelines on community-level interventions to manage declines in intrinsic capacity: World Health Organization; 2017.

8. Agbozo F, Amardi-Mfoafo J, Dwase H, Ellahi B. Nutrition knowledge, dietary patterns and anthropometric indices of older persons in four peri-urban communities in Ga West municipality, Ghana. African health sciences. 2018;18(3):743–55.

9. Blankson B, Hall A. The anthropometric status of elderly women in rural Ghana and factors associated with low body mass index. The Journal of nutrition, health and aging. 2012;16(10):881–6.

10. Aganiba BA, Owusu W, Steiner-Asiedu M, Dittoh S. Association between lifestyle and health variables with nutritional status of the elderly in the northern region of Ghana. African Journal of Food, Agriculture, Nutrition and Development. 2015;15(4):10198–216.

11. Apprey C, Kalog G, Asamoah-Boakye O, Annan R. Nutritional Status and Non-communicable Diseases in Older Ghanaians. 2019.

12. Hayfron ENAA, Annan RA, Edusei AK, Apprey C, Asamoah-Boakye O. Dietary intakes, anthropometric status, and anaemia prevalence among older adults in Effutu Municipality, Ghana. Nutrition and Healthy Aging. 2021;6:49–59.

13. Quesada-Caballero M, Carmona-García A, García-Lara RA, Caballero-Mateos AM, Suleiman-Martos N, Cañadas-De la Fuente GA, et al. Assessing the Risk of Hypertension in Chronic, Elderly Patients during the COVID-19 Pandemic: A Prospective Study. Journal of Cardiovascular Development and Disease. 2024;11(1):21.

14. Dai B, Addai-Dansoh S, Nutakor JA, Osei-Kwakye J, Larnyo E, Oppong S, et al. The prevalence of hypertension and its associated risk factors among older adults in Ghana. Front Cardiovasc Med. 2022;9:990616.

15. Biritwum R, Mensah G, Yawson AE, Minicuci N, Kowal P. Ghana: Study on global AGEing and adult health (SAGE) Wave 1 National Report 2015.

16. Takase M, Takahashi K, Ogino R, Nitanai R, Tanaka T, Saisho S, et al. Functional capacity in community-dwelling older adults maintained by a higher friend network than family network: implications from a two-year longitudinal study. BMC Research Notes. 2022;15(1):319.

17. Puto G, Repka I, Muszalik M. Factors Correlating with Functional Capacity in Older People with Chronic Pain. Int J Environ Res Public Health. 2023;20(3).

18. Baugreet S, Hamill RM, Kerry JP, McCarthy SN. Mitigating Nutrition and Health Deficiencies in Older Adults: A Role for Food Innovation? Journal of Food Science. 2017;82(4):848–55.

19. Agostoni C, Bresson JL, Fairweather Tait S, Flynn A, Golly I, Korhonen H, et al. Scientific opinion on dietary reference values for protein: EFSA panel on dietetic products, nutrition and allergies (NDA). EFSA JOURNAL. 2012;10(2):1–66.

20. Nations. FaAOotU. MINIMUM DIETARY DIVERSITY FOR WOMEN: An Updated Guide to Measurement-from Collection to Action: FOOD & AGRICULTURE ORG; 2021.

21. Alhussain MH, Alkahtani S, Aljuhani O, Habib SS. Effects of Nutrient Intake on Diagnostic Measures of Sarcopenia among Arab Men: A Cross-Sectional Study. Nutrients. 2020;13(1).

22. Pereira SL, Shoemaker ME, Gawel S, Davis GJ, Luo M, Mustad VA, et al. Biomarker Changes in Response to a 12-Week Supplementation of an Oral Nutritional Supplement Enriched with Protein, Vitamin D and HMB in Malnourished Community Dwelling Older Adults with Sarcopenia. Nutrients. 2022;14(6).

23. Beaudart C, Locquet M, Touvier M, Reginster J-Y, Bruyère O. Association between dietary nutrient intake and sarcopenia in the SarcoPhAge study. Aging Clinical and Experimental Research. 2019;31(6):815–24.

24. Park S, Ham J-O, Lee B-K. A positive association of vitamin D deficiency and sarcopenia in 50 year old women, but not men. Clinical Nutrition. 2014;33(5):900–5.

25. Washburn RA, Smith KW, Jette AM, Janney CA. The physical activity scale for the elderly (PASE): Development and evaluation. Journal of Clinical Epidemiology. 1993;46(2):153–62.

26. Menber Y, Gashaw S, Belachew T, Fentahun N. Validation of the minimum dietary diversity for women as a predictor of micronutrient adequacy among lactating women in Ethiopia. Frontiers in Nutrition. 2024;Volume 11 - 2024.

27. Control H. Report of a WHO Expert Committee. WHO Technical Report Series 862. World Health Organization Geneva; 1996.

28. Amfo-Antiri A, Agyapong NAF, Cobbah L. Dietary Habits and Nutritional Challenges of the Elderly in Ghana. J Nutr Metab. 2023;2023:3011067.

29. Oldewage-Theron WH, Samuel FO, Djoulde RD. Serum concentration and dietary intake of vitamins A and E in low-income South African elderly. Clinical Nutrition. 2010;29(1):119–23.

30. Rathnayake KM, Wimalathunga M, Weech M, Jackson KG, Lovegrove JA. High prevalence of undernutrition and low dietary diversity in institutionalised elderly living in Sri Lanka. Public Health Nutr. 2015;18(15):2874–80.

31. Islam N, Islam M, Nahar Q. Assessment of Micro Nutritional Malnutrition in an Urban Area of Bangladesh among the Adult Population on the Basis of Individual Dietary Diversity Score (IDDS). International Journal of Nutrition and Food Sciences. 2015;5:362.

32. Cederholm T, Jensen GL, Correia M, Gonzalez MC, Fukushima R, Higashiguchi T, et al. GLIM criteria for the diagnosis of malnutrition -A consensus report from the global clinical nutrition community. J Cachexia Sarcopenia Muscle. 2019;10(1):207–17.

33. Cruz-Jentoft AJ, Bahat G, Bauer J, Boirie Y, Bruyère O, Cederholm T, et al. Sarcopenia: revised European consensus on definition and diagnosis. Age and ageing. 2019;48(1):16–31.

34. Mezgebu GS, Petros L, Alemayew E, Abebaw G, Feleke FW. Magnitude of undernutrition and its association with dietary diversity among older persons in Ethiopia: a systematic review and meta-analysis, 2023. J Nutr Sci. 2023;12:e102.

35. Vaishya R, Misra A, Vaish A, Ursino N, D’Ambrosi R. Hand grip strength as a proposed new vital sign of health: a narrative review of evidences. Journal of Health, Population and Nutrition. 2024;43(1):7.

36. Koopman JJE, van Bodegom D, van Heemst D, Westendorp RGJ. Handgrip strength, ageing and mortality in rural Africa. Age and Ageing. 2014;44(3):465–70.

37. Arokiasamy P, Selvamani Y, Jotheeswaran AT, Sadana R. Socioeconomic differences in handgrip strength and its association with measures of intrinsic capacity among older adults in six middle-income countries. Sci Rep. 2021;11(1):19494.

38. Wearing J, Konings P, Stokes M, de Bruin ED. Handgrip strength in old and oldest old Swiss adults – a cross-sectional study. BMC Geriatrics. 2018;18(1):266.

39. Kim SH, Hurh K, Park Y, Jang S-I, Park E-C. Synergistic associations of visual and self-reported hearing acuity with low handgrip strength in older adults: a population-based cross-sectional study. BMC Geriatrics. 2021;21(1):513.

40. Doyev R, Axelrod R, Keinan-Boker L, Shimony T, Goldsmith R, Nitsan L, et al. Energy Intake Is Highly Associated with Handgrip Strength in Community-Dwelling Elderly Adults. The Journal of Nutrition. 2021;151(5):1249–55.

41. Lloyd-Jones DM, Evans JC, Levy D. Hypertension in adults across the age spectrum: current outcomes and control in the community. Jama. 2005;294(4):466–72.

42. Calys-Tagoe B, Nuertey BD, Tetteh J, Yawson AE. Individual awareness and treatment effectiveness of hypertension among older adults in Ghana: evidence from the World Health Organization study of global ageing and adult health wave 2. Pan Afr Med J. 2020;37:264.

43. Kim J-H. Age-Specific Association Between Handgrip Strength and Nutritional Quality in Korean Men: A Nationwide Cross-Sectional Study. American Journal of Men’s Health. 2021;15:155798832110633.

44. van Dronkelaar C, van Velzen A, Abdelrazek M, van der Steen A, Weijs PJM, Tieland M. Minerals and Sarcopenia; The Role of Calcium, Iron, Magnesium, Phosphorus, Potassium, Selenium, Sodium, and Zinc on Muscle Mass, Muscle Strength, and Physical Performance in Older Adults: A Systematic Review. Journal of the American Medical Directors Association. 2018;19(1):6–11.e3.

45. Yokoyama Y, Nishi M, Murayama H, Amano H, Taniguchi Y, Nofuji Y, et al. Dietary variety and decline in lean mass and physical performance in community-dwelling older Japanese: A 4-year follow-up study. The Journal of nutrition, health and aging. 2017;21(1):11–6.

46. Otsuka R, Kato Y, Nishita Y, Tange C, Nakamoto M, Tomida M, et al. Dietary diversity and 14-year decline in higher-level functional capacity among middle-aged and elderly Japanese. Nutrition. 2016;32(7):784–9.

47. Landi F, Calvani R, Tosato M, Martone AM, Ortolani E, Savera G, et al. Protein Intake and Muscle Health in Old Age: From Biological Plausibility to Clinical Evidence. Nutrients. 2016;8(5):295.

48. Smith GI, Julliand S, Reeds DN, Sinacore DR, Klein S, Mittendorfer B. Fish oil– derived n−3 PUFA therapy increases muscle mass and function in healthy older adults1. The American Journal of Clinical Nutrition. 2015;102(1):115–22.

49. Kim MK, Baek KH, Song K-H, Il Kang M, Park CY, Lee WY, et al. Vitamin D Deficiency Is Associated with Sarcopenia in Older Koreans, Regardless of Obesity: The Fourth Korea National Health and Nutrition Examination Surveys (KNHANES IV) 2009. The Journal of Clinical Endocrinology & Metabolism. 2011;96(10):3250–6.

50. Kim MH, Choi MK, Bae YJ. Relationship between protein intake and grip strength in qualitative and quantitative aspects among the elderly in Korea: results from the Korea National Health and Nutrition Examination Survey. BMC Geriatrics. 2023;23(1):330.

51. Tak YJ, Lee JG, Yi YH, Kim YJ, Lee S, Cho BM, et al. Association of Handgrip Strength with Dietary Intake in the Korean Population: Findings Based on the Seventh Korea National Health and Nutrition Examination Survey (KNHANES VII-1), 2016. Nutrients. 2018;10(9).

52. Gi Y-m, Jung B, Kim K-W, Cho J-H, Ha I-H. Low handgrip strength is closely associated with anemia among adults: A cross-sectional study using Korea National Health and Nutrition Examination Survey (KNHANES). PLOS ONE. 2020;15:e0218058.

53. Kim BM, Yi YH, Kim YJ, Lee SY, Lee JG, Cho YH, et al. Association between Relative Handgrip Strength and Dyslipidemia in Korean Adults: Findings of the 2014-2015 Korea National Health and Nutrition Examination Survey. Korean J Fam Med. 2020;41(6):404–11.

54. Lee Y, Lim J-Y, Lee S. Relationship Between Low Handgrip Strength and Chronic Kidney Disease: KNHANES 2014-2017. Journal of Renal Nutrition. 2020;31.

55. Ji C, Zheng L, Zhang R, Wu Q, Zhao Y. Handgrip strength is positively related to blood pressure and hypertension risk: Results from the National Health and nutrition examination survey. Lipids in Health and Disease. 2018;17.

56. Hong S. Association of Relative Handgrip Strength and Metabolic Syndrome in Korean Older Adults: Korea National Health and Nutrition Examination Survey VII-1. JOMES. 2019;28(1):53–60.

57. Kang Y, Kim J, Kim DY, Kim S, Park S, Lim H, et al. Association between Dietary Patterns and Handgrip Strength: Analysis of the Korean National Health and Nutrition Examination Survey Data Between 2014 and 2017. Nutrients. 2020;12(10).

58. Ribeiro LW, Berndt S, Mielke GI, Doust J, Mishra GD. Factors associated with handgrip strength across the life course: A systematic review. J Cachexia Sarcopenia Muscle. 2024;15(6):2270–80.

59. Huang J, Shanmugam A, Huang X, van Dam R, Hilal S. Association of Diet Quality with Hand Grip Strength Weakness and Asymmetry in A Multi-ethnic Asian Cohort. The British journal of nutrition. 2023;131:1–21.

60. Bahadoran Z, Golzarand M, Mirmiran P, Shiva N, Azizi F. Dietary total antioxidant capacity and the occurrence of metabolic syndrome and its components after a 3-year follow-up in adults: Tehran Lipid and Glucose Study. Nutrition & Metabolism. 2012;9(1):70.

61. De la Iglesia R, Lopez-Legarrea P, Celada P, Sánchez-Muniz FJ, Martinez JA, Zulet MA. Beneficial Effects of the RESMENA Dietary Pattern on Oxidative Stress in Patients Suffering from Metabolic Syndrome with Hyperglycemia Are Associated to Dietary TAC and Fruit Consumption. International Journal of Molecular Sciences. 2013;14(4):6903–19.

62. Moreira SR, Cucato GG, Terra DF, Ritti-Dias RM. Acute blood pressure changes are related to chronic effects of resistance exercise in medicated hypertensives elderly women. Clin Physiol Funct Imaging. 2016;36(3):242–8.

63. Distefano G, Goodpaster BH. Effects of Exercise and Aging on Skeletal Muscle. Cold Spring Harb Perspect Med. 2018;8(3).

64. Graham ZA, Lavin KM, O’Bryan SM, Thalacker-Mercer AE, Buford TW, Ford KM, et al. Mechanisms of exercise as a preventative measure to muscle wasting. American Journal of Physiology-Cell Physiology. 2021;321(1):C40–C57.

